# Gut inflammation associated with age and Alzheimer’s disease pathology

**DOI:** 10.1101/2022.09.21.22280179

**Authors:** Margo B. Heston, Kendra L. Hanslik, Katie R. Zarbock, Sandra J. Harding, Nancy J. Davenport-Sis, Robert L. Kerby, Nathaniel Chin, Yi Sun, Ana Hoeft, Yuetiva Deming, Nicholas M. Vogt, Tobey J. Betthauser, Sterling C. Johnson, Sanjay Asthana, Gwendlyn Kollmorgen, Ivonne Suridjan, Norbert Wild, Henrik Zetterberg, Kaj Blennow, Federico E. Rey, Barbara B. Bendlin, Tyler K. Ulland

**Affiliations:** Wisconsin Alzheimer’s Disease Research Center, University of Wisconsin School of Medicine and Public Health; Madison, Wisconsin, USA; Department of Pathology and Laboratory Medicine, University of Wisconsin-Madison; Madison, Wisconsin, USA; Department of Bacteriology, University of Wisconsin-Madison; Madison, Wisconsin, USA; Wisconsin Alzheimer’s Institute, University of Wisconsin School of Medicine and Public Health; Madison, Wisconsin, USA; Roche Diagnostics GmbH; Penzberg, Germany; Roche Diagnostics International Ltd; Rotkreuz, Switzerland; Department of Psychiatry and Neurochemistry, Institute of Neuroscience and Physiology, the Sahlgrenska Academy at the University of Gothenburg; Mölndal, Sweden; Clinical Neurochemistry Laboratory, Sahlgrenska University Hospital; Mölndal, Sweden; Department of Neurodegenerative Disease, UCL Institute of Neurology; London, UK; UK Dementia Research Institute at UCL; London, UK; Hong Kong Center for Neurodegenerative Diseases; Hong Kong, China

## Abstract

Age-related disease may be mediated by low levels of chronic inflammation (“inflammaging”). Recent work suggests that gut microbes may contribute to inflammation via degradation of the intestinal barrier. While aging and age-related diseases including Alzheimer’s disease (AD) are linked to altered microbiome composition and higher levels of gut microbial components in systemic circulation, the role of intestinal inflammation and permeability *per se* remains unclear. To test whether greater gut inflammation is associated with older age and AD pathology, we assessed fecal samples from older adults to measure calprotectin, an established marker of intestinal inflammation which is elevated in diseases of gut barrier integrity. Here we found that calprotectin levels are higher with age, and that higher calprotectin was associated with greater amyloid burden among participants with an amyloid-confirmed AD dementia diagnosis. Calprotectin was also associated with cerebrospinal fluid markers of AD pathology and axonal degeneration, as well as with lower verbal memory function among cognitively unimpaired participants. Together, these findings suggest that intestinal inflammation may play a role in pathology development, and that it may exacerbate the progression toward AD.

**Summary:** Intestinal inflammation is correlated with older age, Alzheimer’s disease (AD) dementia, and greater amyloid burden in participants with AD.

## INTRODUCTION

Inflammaging is a hallmark of age that is characterized by chronic low-grade systemic inflammation without an acute driver such as infection *(1)*. Proposed by Franceschi and colleagues as an immune response to continuous environmental and antigenic stressors in age, inflammaging is correlated with frailty and decline across multiple organ systems, age-related diseases including diabetes, cancer, and neurodegenerative diseases, and higher rates of all-cause mortality *(1–6)*. Many stressors that drive inflammaging are unknown, as is the mechanistic role that inflammaging serves in age-related disease *(3)*. It is particularly unclear whether age-associated inflammation precedes or results from the chronic conditions it frequently accompanies. As age-related diseases – many of which prominently feature inflammation – represent over 50% of global disease burden in adults, the drivers of inflammaging and its role in disease require further research emphasis *(7–10)*.

The gut microbiota is hypothesized to contribute to inflammaging, yet the physiological mechanisms by which this occurs remain underexplored *(11–14)*. It is possible that age-related shifts in microbiome composition produce an inflammatory environment within the intestine, which degrades the epithelial barrier. This may enable luminal bacterial components such as lipopolysaccharides and peptidoglycan fragments to enter systemic circulation, eliciting a peripheral inflammatory response *(11, 15–17)*. Evidence suggests that gut permeability increases with aging and age-related diseases. For instance, levels of high-mobility group box 1 protein, which induces intestinal inflammation, are correlated with advanced age as well as markers of chronic inflammation, frailty, and impaired nutrient absorption *(18–20)*. Further, elevated permeability appears more frequently among older adults with inflammatory conditions including type 2 diabetes and irritable bowel syndrome (IBS) *(11, 21)*. However, the few studies of intestinal permeability in healthy aging adults have yielded inconclusive evidence, so it remains unclear whether elevated inflammation and permeability can occur prior to the onset of age-related disease.

Among neurodegenerative diseases, mounting evidence suggests that gut dysbiosis and peripheral inflammation are associated with Alzheimer’s disease (AD) pathology and may contribute to AD pathogenesis *(5, 6, 22–24)*. AD – which is defined by cerebral aggregation of amyloid beta (Aβ) and phosphorylated tau (pTau) proteins – is accompanied by microglial and astrocytic activation and associated neuroinflammation *(25–28)*. Glial activation may exacerbate or mitigate deposition of pathogenic Aβ and pTau, depending on disease stage *(26, 27)*. While some glial activation triggers have been established, including Aβ, microglial activators are diverse and underexplored. Candidate activators may include microbial-derived molecules including metabolites and toxins in systemic circulation, as microglia respond to numerous damage- and microbe-associated molecular patterns arising during cellular injury or infection *(29, 30)*. Furthermore, factors including gut dysbiosis and gut bacteria derived metabolites have been linked to inflammation, preclinical cognitive decline, and AD diagnosis, but their mechanistic relationships with pathology remain ill-defined *(23, 24, 31–35)*. Interestingly, a nationwide population study in Taiwan (1,742 cases, 17,420 controls) found that inflammatory bowel disease (IBD) was associated with higher risk of developing AD dementia, suggesting a possible role for intestinal inflammation and permeability in AD *(36)*. Together, these prior observations, as well as gaps in current knowledge, motivated the current evaluation of intestinal inflammation in aging and as a precursor to AD pathology.

To index gut inflammation, we collected fecal samples from middle and older-aged adults along the biologic and clinical AD continuum and measured fecal calprotectin, a marker of intestinal inflammation. Calprotectin is a heterodimer of S100 calcium-binding proteins expressed primarily in myeloid cells and released from the cytosol of activated neutrophils *(37, 38)*. This molecular complex is suitable as a biomarker due to its abundance in neutrophils, constituting approximately 60% of their cytosolic protein, and its stability at room temperature for up to 7 days *(39–41)*. Calprotectin is also a marker specific to inflammation, as several *in vitro* and clinical studies indicate that calprotectin plays a role in monocyte recruitment *(42–44)*. In diagnostic practice, fecal calprotectin has shown utility as a marker of intestinal inflammation. Notably, fecal calprotectin distinguishes between IBD, which is marked by increased gut permeability, from functional gastrointestinal disorders *(45–47)*. Together, fecal calprotectin’s abundance, stability, and specificity position it as a strong biomarker of intestinal permeability.

To probe the effects of intestinal inflammation on age and AD, we tested associations of fecal calprotectin with years of age, cognitive function, and markers of AD pathology indexed by positron emission tomography (PET) neuroimaging and cerebrospinal fluid (CSF) biomarkers. We hypothesized that prior to symptomatic AD onset, older age would be associated with greater fecal calprotectin. We also hypothesized that higher calprotectin would be associated with greater regional Aβ burden in the brain, higher concentration of CSF biomarkers of AD pathology, glial activation, and neurodegeneration, and lower cognitive function. Further, given that the effects of inflammation may have different relationships with AD pathology dependent on disease stage, we hypothesized that calprotectin effects on AD outcomes would vary across the AD continuum; we tested this using a calprotectin-by-stage interaction effect. To test these hypotheses, we obtained measurements from participants across the AD continuum, and classified three disease stages using clinical diagnosis and amyloid positivity: cognitively unimpaired and Aβ-negative (CU Aβ-), cognitively unimpaired and Aβ-positive (CU Aβ+), and AD dementia with confirmed Aβ positivity (AD dementia). Linear regression was used to test the extent to which intestinal inflammation was associated with age, cerebral amyloid retention, and cognitive decline.

## RESULTS

### Participant demographics

125 participants (CU Aβ-, n=79; CU Aβ+, n=33; AD dementia, n=13) provided samples which were assessed for levels of fecal calprotectin. Presence or absence of AD dementia was determined in accordance with the 2011 National Institute on Aging Alzheimer’s Association (NIA-AA) workgroup diagnostic criteria and reviewed at a diagnosis consensus meeting. Amyloid status was determined via CSF collected at a lumbar puncture visit or neuroimaging visit closest to the date of fecal sample collection. Amyloid positivity was indexed by at least one of three possible Aβ burden measures: global ^11^C-Pittsburgh compound B (PiB) PET distribution volume ratio (DVR) greater than 1.19, [^18^F]AV45 PET positivity assessed by visual inspection by an expert rater (SCJ), or CSF Aβ_42_/Aβ_40_ ratio less than 0.046 *(48, 49)*. The CSF Aβ_42_/Aβ_40_ threshold was established to maximize agreement between CSF and PET measures of amyloid positivity. Participants with a banked fecal sample, consensus diagnosis, and amyloid burden confirmed within 2 years of a fecal sample were included in the study.

Across biologic and clinical AD stages, participant groups did not significantly differ in sex composition or mean body mass index (BMI), and fecal samples did not differ in mean Bristol Stool Scale (BSS) score, a measure of stool consistency. Educational attainment was generally high, however participants with AD dementia had comparably lower years of education (P=0.054). CU Aβ-participants had completed a mean ± standard deviation (SD) 16.41 ± 2.71 years of education and CU Aβ+ participants had completed 16.18 ± 2.26 years (equivalent to a Bachelor’s degree), while AD dementia participants had completed 14.69 ± 2.02 years (Associate’s degree). Participants with AD dementia were older on average (mean ± SD: 73.9 ± 5.09 years) compared with CU Aβ-(65.92 ± 5.97 years) and CU Aβ+ (68.92 ± 6.34 years) participants. CU Aβ+ and AD dementia participants were enriched for *APOE* ε4 carriage (percent carriage: 54.5% and 84.6%, respectively) compared with CU Aβ-participants (26.6%). Table 1 describes the demographics for the full cohort, and tables S1-S4 feature the demographics for each subset in analyses of PiB PET, CSF biomarkers, and neuropsychological assessment as outcomes. Each participant subset was compositionally representative of the full cohort.

**Table 1.**
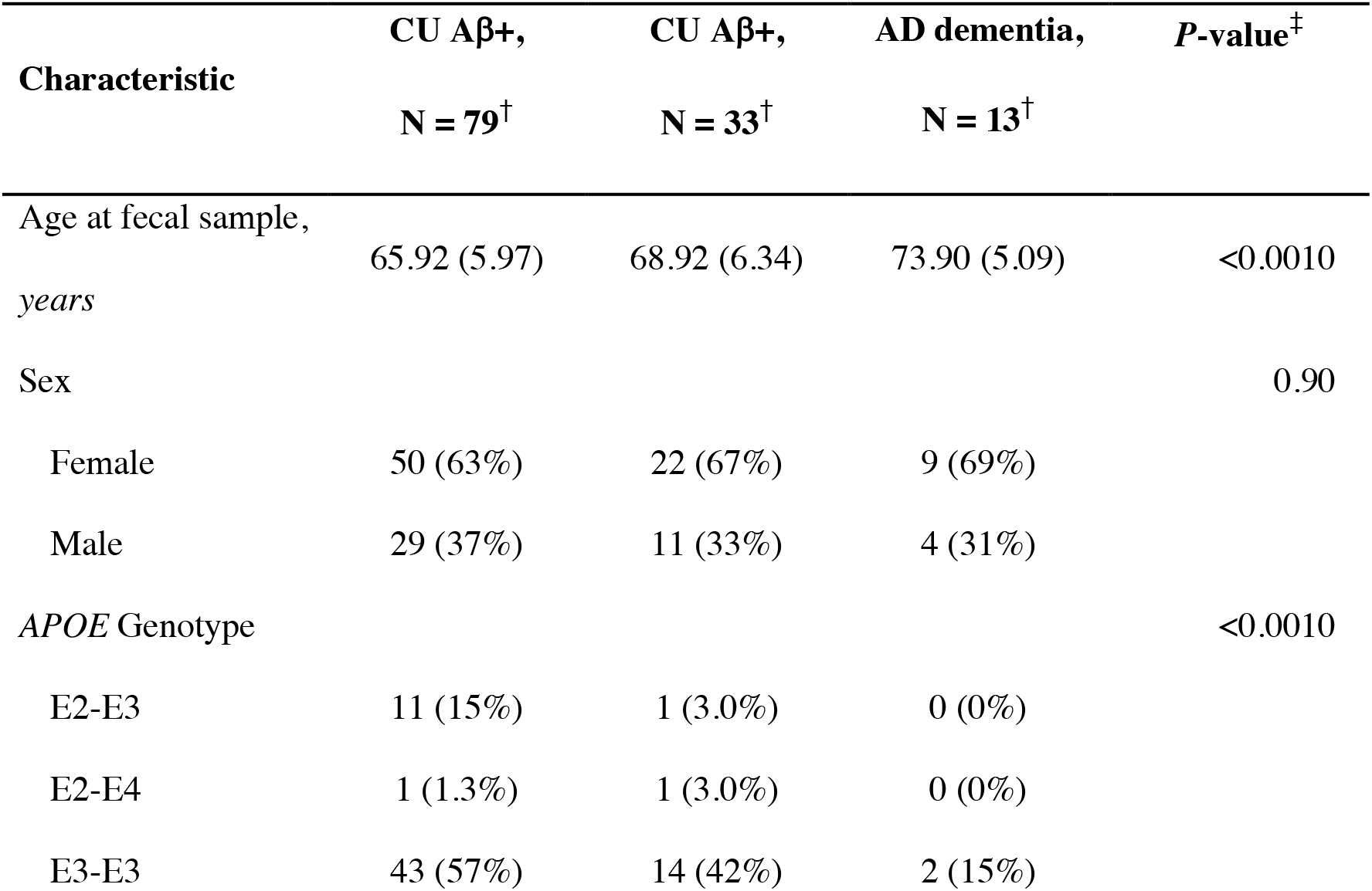
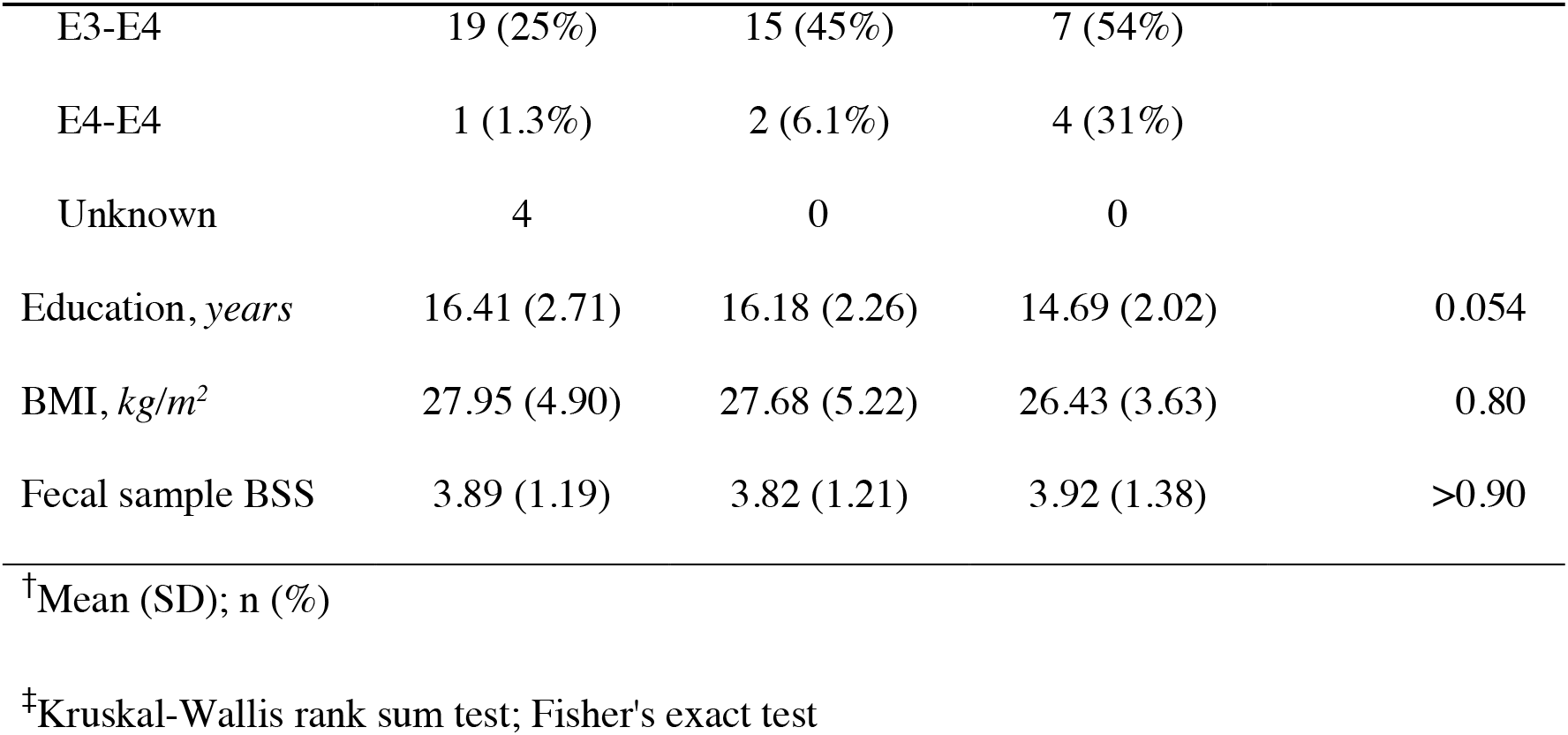
Participant Demographics. Bristol Stool Scale (BSS) ranges from 1 (hard lumps) to 7 (liquid).

### Calprotectin is higher in older participants and those with AD dementia

To test the hypothesis that intestinal inflammation increases with age and AD progression, fecal calprotectin was regressed on participant age at fecal sample, and on disease stage (CU Aβ-, CU Aβ+, AD dementia). Across the full cohort, older age was significantly associated with greater calprotectin (*β* [95% CI] = 0.016 [0.002, 0.03], P=0.027), controlling for effects of disease stage, sex, and BMI (**Fig. 1A**). This effect was sustained upon restricting analysis to participants with a CU consensus diagnosis (0.015 [6.0e-4, 0.03], P=0.042) (**Fig. 1B**). Regression effects for the full cohort are tabulated in table S5, and effects for CU-only participants are tabulated in table S6. In addition, mean calprotectin was significantly higher among AD dementia participants compared with CU Aβ-participants (0.28 [0.08, 0.49], P=0.010) and with CU Aβ+ participants (0.24 [0.02, 0.47], P=0.030) (**Fig. 2**).

**Fig. 1.**
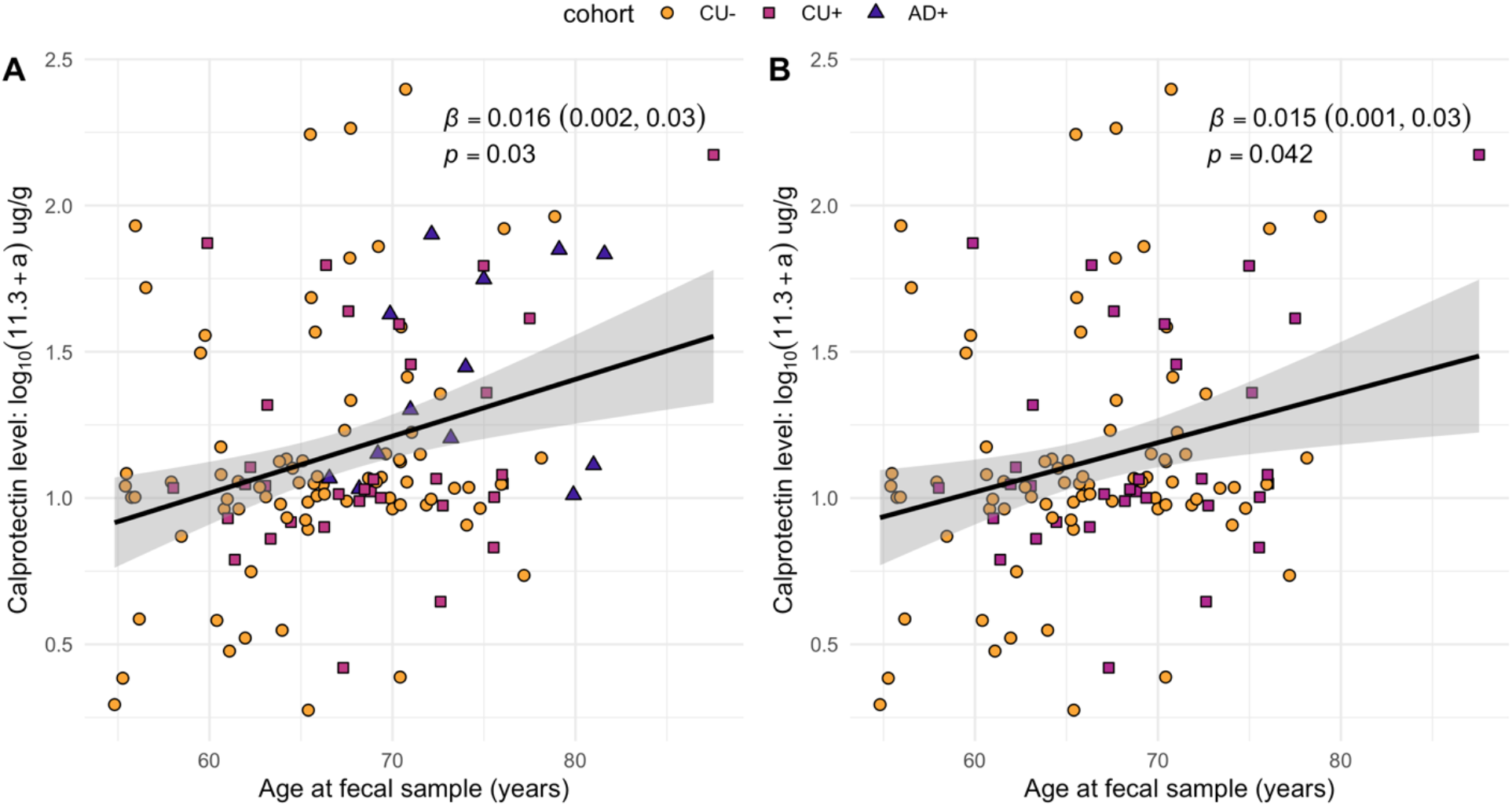
Calprotectin association with age. **(A**) Relationship between participant age and fecal calprotectin levels across all disease statuses. **(B**) Relationship between participant age and fecal calprotectin levels before cognitive decline. β coefficients (multiple regression) were controlled for cohort, sex, and BMI.

**Fig. 2.**
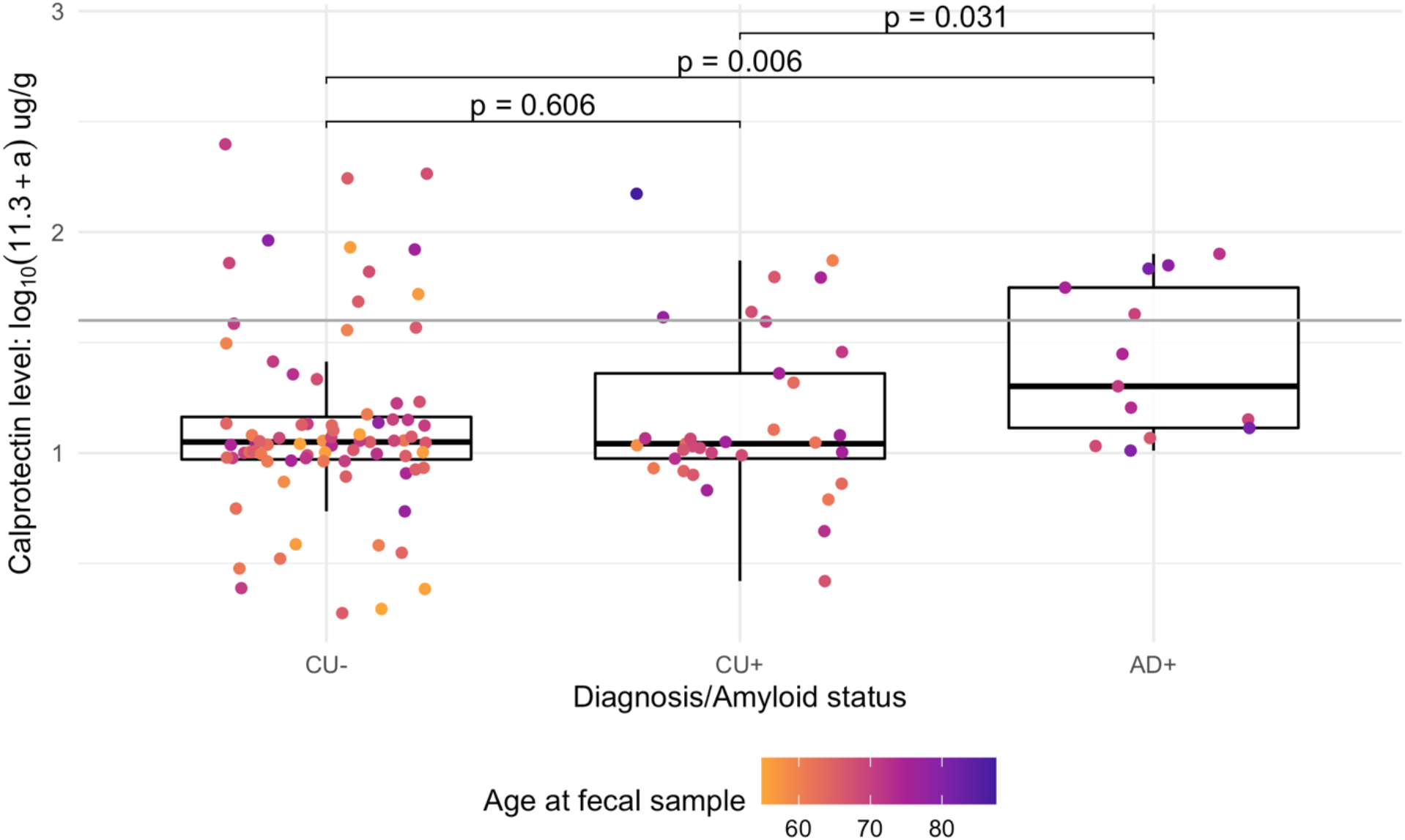
Calprotectin association with disease status. Fecal calprotectin is considered elevated at 43.2 µg/g or 1.6 log(µg/g), denoted by the gray horizontal line and defined by the ELISA standard curve. *P*-values (multiple regression) are uncontrolled for covariates.

While age and AD dementia status were associated with greater calprotectin, visual inspection of calprotectin plots revealed that several CU Aβ-participants (n=10) exhibited elevated calprotectin relative to the high-calprotectin threshold of the enzyme-linked immunosorbent assay (ELISA), which was not significantly associated with older age (P=0.40, table S7). To identify factors that distinguished the high-calprotectin CU Aβ-participants from their low-calprotectin counterparts, a *post hoc* analysis was conducted on self-reported comorbidities, medications, dietary patterns, and other microbiome-altering lifestyle factors *(50– 52)*. Of the factors examined, there were no differences in prevalence of reported IBD, IBS, and probiotic usage. Only proton pump inhibitor (PPI) usage was identified as being more frequent among high-calprotectin CU-participants (P=0.013, table S7), with 4 in 10 of these participants reporting use of PPI.

### Calprotectin-by-AD dementia interaction on PiB cortical DVR

The main effects of gut inflammation and the interaction between gut inflammation and disease status on brain Aβ burden were tested using PiB PET data. PiB DVR in 8 cortical regions known to accumulate pathology were assessed separately as well as averaged to derive a global measure. Regions included the anterior cingulate cortex (ACC), angular gyrus (ANG), middle temporal gyrus (MTG), posterior cingulate cortex (PCC), precuneus (PCUN), supramarginal gyrus (SMG), superior temporal gyrus (STG), and ventromedial prefrontal cortex (vmPFC).

Regression analysis tested the main effects as well as a calprotectin-by-disease status interaction. Eighty-seven individuals with fecal calprotectin, disease status, and PiB PET neuroimaging collected within 2 years of a fecal sample were included. Globally and within the ACC, PCC, vmPFC, PCUN, and MTG regions, PiB cortical DVR was positively associated with calprotectin within the AD dementia participant subset (**Fig. 3**). Except in the MTG, all effects remained significant after adjustment for age, and effects in the PCC and PCUN remained significant after correction for age, sex, APOE risk score and BMI (table S8). CU Aβ+ did not show a significant relationship between calprotectin and PiB cortical DVR in any regions or with global DVR (table S9). To test the extent to which disease stage interactions were confounded by age, PiB PET models were re-evaluated using a calprotectin-by-age interaction term. Prior to controlling for covariates, all calprotectin-by-age effects were found non-significant, suggesting a pathology-specific effect (table S10).

**Fig. 3.**
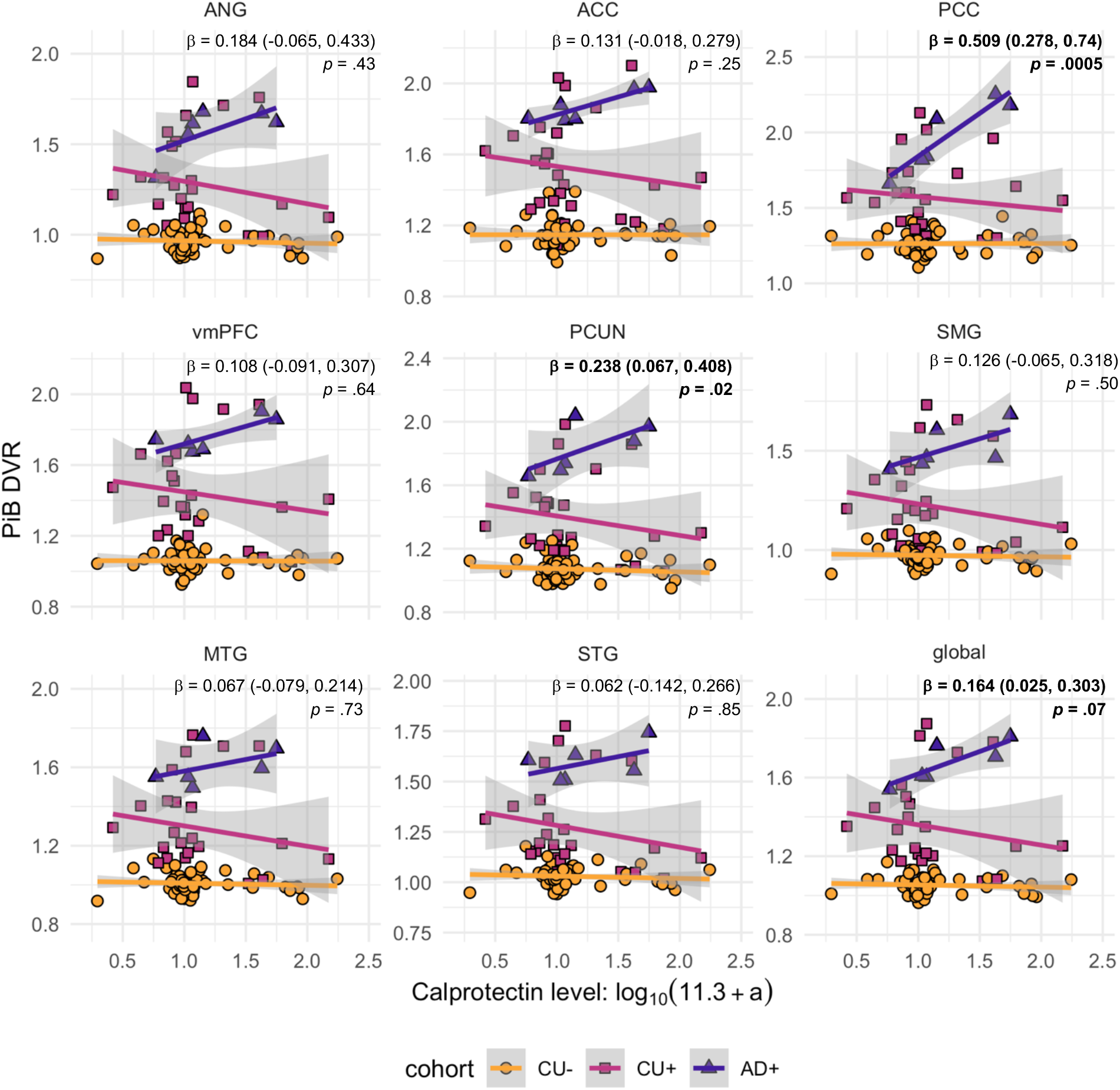
Calprotectin-by-AD dementia interaction effects on regional PiB cortical DVR. β coefficients and *q*-values (multiple regression, false discovery rate-corrected) report the estimates for calprotectin-by-AD dementia interaction effects. Models included calprotectin, cohort, calprotectin-by-cohort interaction, age, sex, *APOE* risk score, and BMI terms.

### CSF biomarkers and cognitive function do not vary significantly with calprotectin level

We conducted exploratory analyses of calprotectin and CSF biomarkers of AD, glial activation, and neurodegeneration *(53)*. Participants with CSF biomarkers within 2 years of a fecal sample were included and 9 CSF biomarkers were separately regressed on fecal calprotectin. Biomarkers included those related to AD pathology (Aβ42/Aβ40, pTau_181_/Aβ42), glial activation (chitinase-3-like protein 1 [YKL-40], glial fibrillary acidic protein [GFAP], soluble triggering receptor expressed on myeloid cells 2 [sTREM2], and S100 calcium binding protein B [S100]), axonal degeneration (neurofilament light [NfL]), and synaptic degeneration (neurogranin, α-synuclein). Prior to adding covariates, higher calprotectin was significantly associated with lower Aβ_42_/Aβ_40_, higher pTau_181_/Aβ42, and higher NfL concentration (P=0.013, P=0.021, and P=0.030, respectively) (**Fig. 4**). However, these effects were not significant after correction for multiple comparisons or adjustment for participant age and disease stage (table S11).

**Fig. 4.**
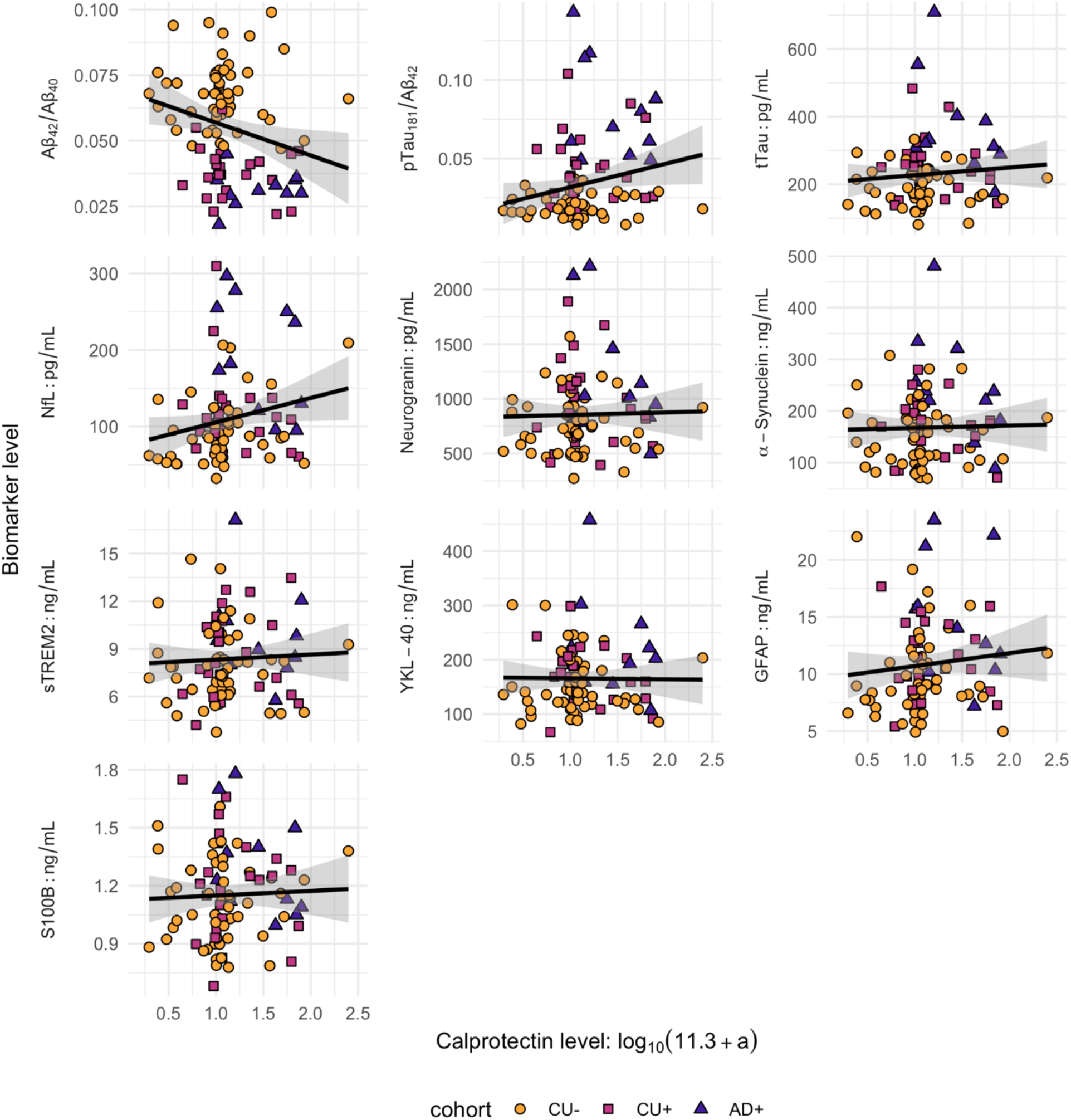
CSF biomarker links to fecal calprotectin. β coefficients and *p*-values (multiple regression) were estimated without controlling for covariates; effects did not survive false discovery rate correction.

To test the hypothesis that intestinal inflammation is associated with lower cognitive function before diagnosis of AD dementia, neuropsychological assessments of verbal learning and executive function were separately regressed on fecal calprotectin. The Rey Auditory Verbal Learning Test (RAVLT) trials 1-5 measured the total words recalled after 5 trials of learning 15 words, and the RAVLT trial 7 measured the total words recalled from this list after a delay. One hundred and three CU participants with cognitive assessments within 2 years of a fecal sample were included. Higher calprotectin was associated with lower delayed recall (fewer words remembered in RAVLT trial 7), controlling for educational attainment and amyloid status. The Trail Making Test (TMT) part B was used to index executive function. Effects of calprotectin on TMT performance were not significant after controlling for age and multiple comparisons adjustment (fig. S1).

## DISCUSSION

Inflammaging is a chronic, low-grade inflammation associated with age which may be driven by alterations in the composition of the gut microbiome and by a reduced selectivity of the intestinal barrier *(3)*. Prior evidence suggests that age-related gut microbiome disruptions may elicit intestinal and systemic inflammation that contributes to diseases, potentially including neurodegenerative diseases such as AD, however the relationships between age, gut permeability and disease markers have not been well established *(3, 23, 24, 34)*. To evaluate the extent to which intestinal inflammation correlates with age and markers of AD, we measured fecal calprotectin levels in CU Aβ-, CU Aβ+, and AD dementia individuals, and found that intestinal inflammation was associated with age, AD dementia status, cortical amyloid among individuals with AD dementia, and CSF biomarkers of AD and neurodegeneration.

While there are few existing human studies on calprotectin, emerging evidence suggests that fecal calprotectin levels may vary across the life course and in age-related disease *(54–58)*. For instance, in a population study of children aged 0 to 18 years old, median fecal calprotectin level among healthy infants under 1 year old was nearly twice that of children between 1 and 18 years old *(59–62)*. One small study showed a 2-to 10-fold difference in fecal calprotectin among 45 healthy participants between 30 and 80 years old, however the vast majority of aging studies have examined calprotectin among cohorts with clinical disease diagnoses *(46, 54, 57, 58, 63)*. While our study was enriched for individuals with biologic and clinical AD, we determined that older participants had higher levels of fecal calprotectin while controlling for disease stage, and the results remained significant after performing sensitivity analyses that included only cognitively unimpaired participants, suggesting that advancing age increases gut inflammation independently of symptomatic AD.

Only one prior study evaluated fecal calprotectin among 22 participants with AD dementia, finding that calprotectin was elevated in this cohort. Similarly, we found that participants in our study with AD dementia had higher calprotectin. Additionally, we leveraged *in vivo* biomarkers including CSF assays and amyloid PET to better determine the relationship between intestinal inflammation and AD pathology. We found that higher calprotectin was associated with greater cortical amyloid burden among AD dementia participants, an effect that was independent of age. Calprotectin also nominally correlated with CSF Aβ_42_/Aβ_40_, pTau_181_/Aβ_42_, and NfL. Both CSF Aβ_42_/Aβ_40_ and pTau_181_/Aβ_42_ are markers specific to AD, with the latter biomarker incorporating both amyloid and pTau pathology. NfL is a marker of axonal degeneration, suggesting that intestinal inflammation is also associated with neurodegeneration.

While it may be the case that intestinal inflammation exacerbates the progression of AD, given that the analyses conducted here were cross-sectional, we cannot rule out the possibility that development of AD pathology may exacerbate intestinal inflammation. However, it is interesting to note that even among cognitively unimpaired participants, we observed a relationship between intestinal inflammation and memory function. Previously, it was observed that cognitive dysfunction was associated with intestinal inflammation in other conditions such as Crohn’s disease and IBS *(64, 65)*. These conditions were excluded from the current study; accordingly, calprotectin levels in our study were generally lower than compared with the levels observed among individuals with these conditions. Taken together with prior studies on IBS and IBD, our results showing relationships between calprotectin and memory function suggest that interventions that mitigate intestinal inflammation could have a beneficial impact on cognitive function in older adults.

In addition to associations of calprotectin with age and AD pathology, the present study also suggested that elevated intestinal inflammation may be present among otherwise healthy adults who are negative for amyloid pathology (CU Aβ-). A greater proportion of high-calprotectin participants reported using PPI medications. PPIs are associated with lower microbiome α-diversity, higher abundances of genera including *Enterococcus*, and lower abundances of genera including *Bifidobacterium (52). Enterococcus faecalis* has been shown to generate neurofibrillary epitopes in rat cortical neurons, while *Bifidobacterium* species are differentially abundant in AD compared with controls *(24, 66, 67). Bifidobacterium* species produce numerous potentially disease-modifying metabolites, including peptides which inhibit angiotensin converting enzyme activity and short-chain fatty acids that contribute to intestinal barrier maintenance*(68, 69)*. Apart from PPI usage, there were no disease-related trends which explained the high calprotectin observed in fecal samples of CU Aβ-participants. Identifying elevated calprotectin among otherwise healthy participants may provide an opportunity to determine associations with later disease development and may reveal points of intervention to restore intestinal barrier function. Future longitudinal studies are needed to determine whether high intestinal inflammation predicts greater pathology and accelerated cognitive decline at subsequent time points.

While the present study has elucidated age and AD relationships with calprotectin, some limitations should be noted. The research cohorts included in this study are lacking in diversity (participants were predominantly White and highly educated) and may not reflect the broader population. Continuous sampling of the enrolled individuals will enable longitudinal studies of gut inflammation effects on AD, and active efforts to recruit community members will enable futures studies of the intestinal barrier in a more diverse aging population. Aside from sample limitations, while measuring fecal calprotectin enabled us to capture inflammation in the large intestine, the effects of the small intestine on aging and AD pathology remain unknown. Emerging evidence from human studies suggests that the small intestine exhibits microbial alterations in aging and AD, and increased permeability with age. For instance, a study evaluating the duodenal microbiome of individuals from 18 to 80 years old noted a reduction of microbial diversity and an increase of *Escherichia, Lactobacillus*, and *Enterococcus* genera with age *(70)*. Among a small cohort, individuals with AD were found to have higher rates of small intestinal bacterial overgrowth, which is interrelated with IBS and hypothesized to induce intestinal permeability via microbial dysbiosis *(21, 58, 71, 72)*. Separately, reductions in the ionic gradient of the small intestine have been noted with age *(21)*. In future studies we aim to examine small intestinal permeability and gut microbiome composition as it relates to age and preclinical AD pathology.

In summary, our results suggest that aging may co-occur with intestinal inflammation, that intestinal inflammation is observed in AD dementia and may exacerbate amyloid accumulation, and that associations with cognitive function may appear even prior to development of symptomatic AD dementia. Together, these results suggest intestinal inflammation and possible accompanying intestinal permeability could be a modifiable target in aging and AD.

## MATERIALS AND METHODS

### Recruitment and clinical methods

Participants were recruited from the Wisconsin Alzheimer’s Disease Research Center (ADRC) Clinical Core and the Wisconsin Registry for Alzheimer’s Prevention (WRAP) *(73)*. These prospective cohort studies enroll adults starting in middle age, and cohorts are enriched for participants with parental history of AD. Participants undergo comprehensive cognitive evaluation, interviews of medical and family history, and genotyping of the *APOE* genotyping. Participants also underwent longitudinal cognitive assessments, and a subset underwent lumbar puncture and/or PiB PET neuroimaging. All participants received laboratory testing including blood draws. Participants in this cohort were additionally co-enrolled in the Microbiome in Alzheimer’s Disease Risk Study (MARS), which collects and analyzes fecal samples to determine the role of the gut microbiota in AD development *(24)*.

Clinical consensus diagnosis of cognitively unimpaired, mild cognitive impairment due to AD, or AD dementia was determined via multidisciplinary consensus panel and NIA-AA criteria for AD *(74, 75)*. BMI was calculated from height and weight measurements. As previously published, *APOE* ε2/ε3/ε4 genotyping was performed at baseline using allele-specific polymerase chain reaction-based assays, and for the present study a pseudo-continuous *APOE* risk score was calculated using estimates derived from neuropathologically confirmed cases in the Alzheimer’s Disease Neuroimaging Initiative *(73, 76)* At the time that fecal samples were obtained, participants completed questionnaires detailing current diet and medications, history of gastrointestinal and cardiovascular disease, and lifestyle factors that alter gut microbiome composition, including early childhood microbial exposures and current exposure to house pets or other animals *(24)*. Prior to admission into the WRAP, Wisconsin ADRC, and MARS studies, participants provided written consent in accordance with the Declaration of Helsinki. All work followed approved protocols of the University of Wisconsin-Madison Institutional Review Board.

### Biomarker measurement using CSF and PiB PET

CSF was acquired using the Wisconsin ADRC CSF Biomarker service procedures, typically collected in the morning to control for diurnal variability *(48)*. Biomarkers were measured using the NeuroToolKit for AD pathology (Aβ_42_/Aβ_40_, pTau_181_/Aβ_42_), glial activation (chitinase-3-like protein 1 [YKL-40], glial fibrillary acidic protein [GFAP], soluble triggering receptor expressed on myeloid cells 2 [sTREM2], S100 calcium binding protein B [S100B]), neurodegeneration (total tau [tTau]), axonal degeneration (neurofilament light protein [NfL]), and synaptic degeneration (neurogranin, α-synuclein). CSF biomarkers were quantified using Elecsys^®^ assays for Aβ_42_, pTau_181_, tTau, and S100B were performed on a cobas e^®^ 601 analyzer, and assays for Aβ_40_, YKL-40, GFAP, sTREM2, NfL, neurogranin, and α-synuclein were performed on a cobas e^®^ 411 analyzer *(48, 73)*.

PiB PET scans were acquired, processed, and quantified using published methods *(49)*. Briefly, participants underwent PET (Siemens EXACT HR+) imaging procedures at the University of Wisconsin-Madison Waisman Center. PiB cortical DVR was estimated using reference Logan graphical analysis, and a global cortical DVR average was calculated using DVR from eight bilateral regions that exhibit Aβ burden early in preclinical AD. Regions were determined using the Automated Anatomical Labelling Atlas 3 and included the ACC, ANG, MTG, PCC, PCUN, SMG, STG, and vmPFC.

### Neuropsychological assessment

Participants underwent the RAVLT to assess verbal learning and memory, and the TMT B to assess executive function *(77, 78)*. Raw scores were used as outcome measures in regression analyses and models controlled for age, sex, educational attainment, and amyloid status. The RAVLT Trials 1-5 score denotes the total words learned over 5 repetitions of a 15-word list, and the RAVLT Trial 7 score denotes the words remembered after a delay. The TMT Trial B score indicates the participant’s time in seconds to complete drawing a continuous trail that alternates between ascending numbered and lettered icons printed on the page.

### Fecal collection, processing, and calprotectin measurement

Fecal samples were collected using published methods *(24)*. Briefly, participants completed home sample collection, returning the samples in insulated containers with frozen gel packs via overnight delivery. Samples were subsequently weighed and scored using the BSS. Representative samples of approximately 2-4 g of fecal material were obtained using two sterile straws; both straws were placed in a 15 ml conical tube and stored at -80°C until processing. A subset of samples remained in their collection receptacles without representative sampling. The concentration of calprotectin was measured using an ELISA (Eagle Biosciences, catalog no. CAL35-K01). For each subject, calprotectin levels were measured using aliquots from two distinct locations within the fecal sample. Samples were read at 450 nm with a reference filter at 620 nm, and a cubic equation was used to form a standard curve and calculate data. Based on the assay’s standard curve, calprotectin levels were considered elevated at 43.2 µg/g (or 1.6 log(µg/g) after linearizing the ELISA absorbance distribution; see *Statistical methods* for details) *(79)*. All tests were conducted in duplicate in a blinded manner.

### Statistical methods

To obtain the average calprotectin level for each participant, mean values were calculated using the two fecal sample aliquots. Mean calprotectin levels subsequently underwent a positive translation of 11.3 µg/g and a log_10_ transformation to linearize the absorbance distribution of the ELISA. Multiple linear regression was used to test the extent to which calprotectin level varied with 1) age and diagnosis/amyloid positivity, 2) regional and global PiB cortical DVR, 3) CSF biomarker levels, and 4) tests of cognitive function among CU participants. Calprotectin-by-disease stage interaction predictors were included for PiB PET models, upon visual inspection of the bivariate relationships between calprotectin and biomarker/cognitive outcomes. One model was used per outcome, for a total of 2 age and disease stage models, 9 PiB PET models, 10 CSF biomarker models, and 3 cognitive testing models. When modeling calprotectin as an outcome, covariates included participant sex and BMI. For models involving PET, CSF or cognitive outcomes, covariates included age, sex, amyloid-confirmed consensus diagnosis, BMI, and *APOE* risk score. Regressions were performed in the R package *lavaan*, using maximum likelihood estimation and Satorra-Bentler corrections to adjust for heterogeneity of model residuals *(80, 81)*. Benjamini-Hochberg adjustments were used to control for inflated Type I error rate; adjusted *p*-values are denoted as *q*-values *(82)*. For effects that were found to be significant (Q<0.05) among the full cohort (N=125), models were re-evaluated to determine whether results remained significant among the CU participant subset (n=112).

To identify factors that were associated with elevated calprotectin among CU Aβ-participants, *post hoc* analyses were performed to compare microbiome-altering lifestyle factors, medications, and early life exposure events. Participants with calprotectin levels greater than 1.6 log(µg/g) were assigned to the high-calprotectin group, and the remaining participants were assigned to the low-calprotectin group. Wilcoxon rank sum and Fisher exact tests were used to test factors across the two calprotectin groups.

## Data Availability

Data collected via the Wisconsin ADRC and WRAP protocols may be requested via an online REDCap request tool, which can be accessed at https://www.adrc.wisc.edu/apply-resources.

## List of Supplementary Materials

Fig S1

Tables S1 to S11

## Acknowledgments

We extend our thanks to the WRAP and Wisconsin ADRC participants for their involvement and making this work possible. We thank the staff and researchers at the University of Wisconsin ADRC, the Wisconsin Alzheimer’s Institute, the Wisconsin Institute for Medical Research, and the Waisman Center for their assistance in study organization, participant recruitment, and data collection. NMV is currently affiliated with Massachusetts General Hospital; Boston, Massachusetts, USA.

## Funding

Wisconsin Partnership Program grant (BBB, FER)

Geriatric Research, Education, and Clinical Center of William S. Middleton Memorial Veterans Hospital (BBB, SCJ)

National Institute on Aging grant R01AG070973 (BBB, FER, TKU)

National Institute on Aging grant R01AG070883

National Institute on Aging grant R01AG037639

National Institute on Aging grant R01AG054059

National Institute on Aging grant P30AG062715 (SCJ)

National Institute on Aging grant R01AG021155 (SCJ)

National Institute on Aging grant R01AG27161 (SCJ)

National Institute of Child Health and Human Development grant U54HD090256

National Institute of General Medical Sciences grant T32GM081061 (MBH)

Clinical and Translational Science Award grant UL1TR000427

Swedish Research Council grant 2018-02532 (HZ)

European Research Council grant 681712 (HZ)

European Research Council grant 101053962 (HZ)

Swedish State Support for Clinical Research grant ALFGBG-71320 (HZ)

Swedish State Support for Clinical Research grant ALFGBG-720931 (HZ)

Alzheimer Drug Discovery Foundation grant 201809-2016862 (HZ)

AD Strategic Fund and Alzheimer’s Association grant ADSF-21-831376-C (HZ)

AD Strategic Fund and Alzheimer’s Association grant ADSF-21-831381-C (HZ)

AD Strategic Fund and Alzheimer’s Association grant ADSF-21-831377-C (HZ)

Olav Thon Foundation (HZ)

Erling-Persson Family Foundation (HZ)

Stiftelsen för Gamla Tjänarinnor grant FO2019-0228 (HZ)

European Union Horizon 2020 research and innovation programme under the Marie Skłodowska-Curie grant agreement No 860197 (HZ)

UK Dementia Research Institute at University College London (HZ)

Swedish Research Council grant 2017-00915 (KB)

Alzheimer Drug Discovery Foundation grant RDAPB-201809-2016615 (KB)

Swedish Alzheimer Foundation AF-742881 (KB)

Hjärnfonden, Sweden grant FO2017-0243 (KB)

Swedish State Support for Clinical Research grant ALFGBG-715986 (KB)

European Union Joint Program for Neurodegenerative Disorders grant JPND2019-466-236 (KB) National Institute of Health grant R01AG068398 (KB)

## Author Contributions

Conceptualization: BBB, FER, TKU, MBH

Data curation: MBH, KLH, KRZ

Statistical analysis: MBH

Funding acquisition: BBB, FER, TKU, SCJ, SA

Investigation: KLH, KRZ, SJH, NJD, RLK, YS, AH, NMV, TBJ, HZ, KB, GK

Methodology: BBB, FER, TKU, MBH, RLK, KRZ, KLH

Project administration: SLH, NJD, BBB, FER, TKU

Resources: GK, IS, NW, YD

Supervision: BBB, FER, TKU, SCJ

Visualization: MBH

Writing – original draft: MBH, KLH, KRZ

Writing – review & editing: All authors.

## Competing interests

Roche Diagnostics International Ltd (Rotkreuz, Switzerland) provided CSF testing equipment and analysis for Drs. Bendlin and Johnson. COBAS, COBAS E, and ELECSYS are trademarks of Roche. The Roche NeuroToolKit is a panel of exploratory prototype assays designed to robustly evaluate biomarkers associated with key pathologic events characteristic of AD and other neurological disorders. It is used for research purposes only and is not approved for clinical use.

MBH declares no competing interests.

KLH declares no competing interests.

KRZ declares no competing interests.

SJH declares no competing interests.

NJD declares no competing interests.

RLK declares no competing interests.

YS declares no competing interests.

AH declares no competing interests.

YD declares no competing interests.

NMV declares no competing interests.

TJB declares no competing interests.

SCJ

GK is a full-time employee of Roche Diagnostics GmbH.

IS is a full-time employee of Roche Diagnostics International Ltd.

NW is a full-time employee of Roche Diagnostics GmbH.

HZ has served at scientific advisory boards and/or as a consultant for Abbvie, Acumen, Alector, ALZPath, Annexon, Apellis, Artery Therapeutics, AZTherapies, CogRx, Denali, Eisai, Nervgen, Novo Nordisk, Passage Bio, Pinteon Therapeutics, Red Abbey Labs, reMYND, Roche, Samumed,

Siemens Healthineers, Triplet Therapeutics, and Wave, has given lectures in symposia sponsored by Cellectricon, Fujirebio, Alzecure, Biogen, and Roche, and is a co-founder of Brain Biomarker Solutions in Gothenburg AB (BBS), which is a part of the GU Ventures Incubator Program (outside submitted work).

KB has served as a consultant, at advisory boards, or at data monitoring committees for Abcam, Axon, Biogen, JOMDD/Shimadzu. Julius Clinical, Lilly, MagQu, Novartis, Prothena, Roche Diagnostics, and Siemens Healthineers, and is a co-founder of Brain Biomarker Solutions in Gothenburg AB (BBS), which is a part of the GU Ventures Incubator Program. GK is a full-time employee of Roche Diagnostics GmbH (Penzberg, Germany). IS is a full-time employee and shareholder of Roche Diagnostics International Ltd (Rotkreuz, Switzerland).

BBB has received precursor and imaging agents from AVID radiopharmaceuticals.

FER declares no competing interests.

TKU declares no competing interests.

## Data and materials availability

| **Biomarker** | **Estimate** | **P-value** | **Q-value** |
| --- | --- | --- | --- |
| *Aβ<sub>42</sub>/Aβ<sub>40</sub>* | *-0.013, (-0.022, -0.003)* | *0.013* | *0.10* |
| *pTau<sub>181</sub>/Aβ<sub>42</sub>* | *0.015, (0.002, 0.027)* | *0.021* | *0.10* |
| tTau, pg/mL | 22.763, (-16.774, 62.301) | 0.26 | 0.65 |
| *NFL, pg/mL* | *31.915, (3.017, 60.812)* | *0.030* | *0.10* |
| Neurogranin, pg/mL | 22.361, (-115.105, 159.828) | 0.75 | 0.86 |
| α-Synuclein, ng/mL | 4.521, (-26.288, 35.331) | 0.77 | 0.86 |
| sTREM2, ng/mL | 0.319, (-0.995, 1.634) | 0.63 | 0.86 |
| YKL-40, ng/mL | -1.769, (-34.484, 30.946) | 0.92 | 0.92 |
| GFAP, ng/mL | 1.129, (-1.302, 3.561) | 0.36 | 0.73 |
| S100B, ng/mL | 0.024, (-0.095, 0.143) | 0.69 | 0.86 |

