## Supplementary Figures and Tables for "Gut inflammation associated with age and Alzheimer’s disease pathology"

**Supplementary Materials**

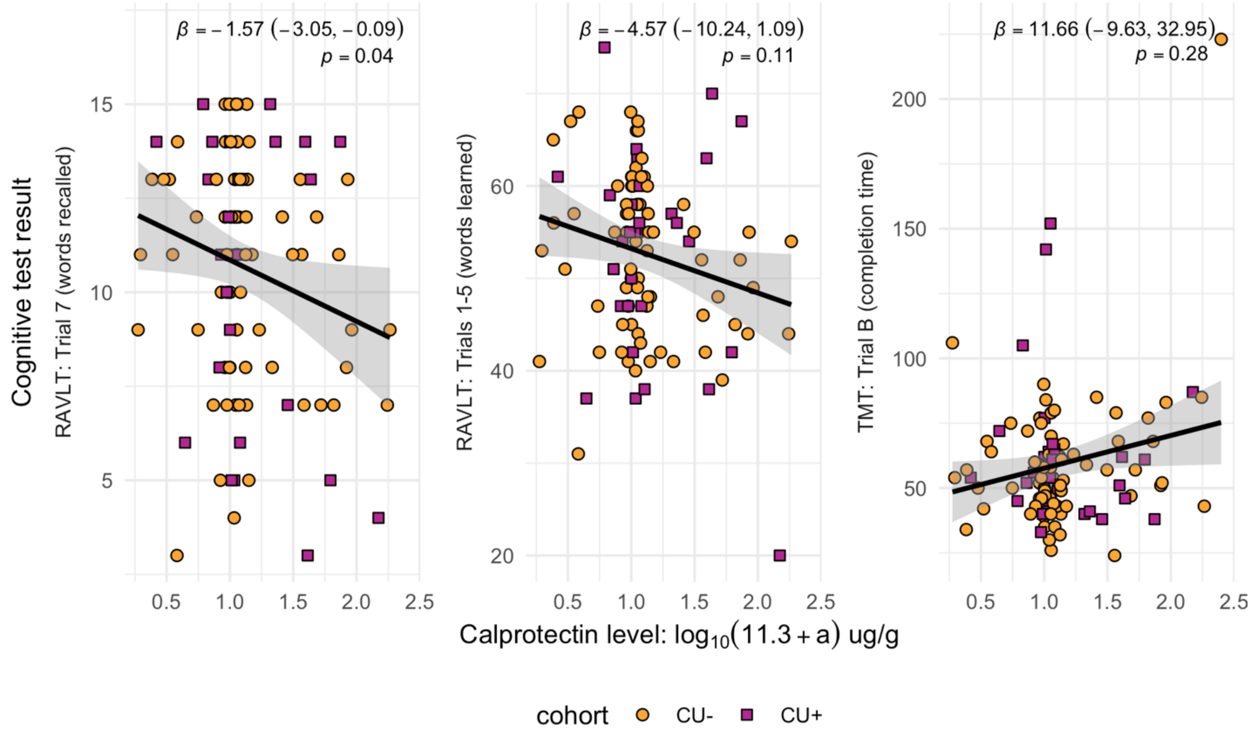


**Fig. S1. Calprotectin effects on cognitive performance in cognitively unimpaired participants.** *CU Aβ-*, cognitively unimpaired, Aβ-negative; *CU Aβ+*, cognitively unimpaired, Aβ-positive; *RAVLT*, Rey Auditory Verbal Learning Test; *TMT*, Trail Making Test. β coefficients (multiple regression, *p*-values false discovery rate uncorrected) are reported with adjustment for education and disease status; no effects survived adjustment for age or sex covariates. An outlier was noted among the TMT Trial B; removal of outlier did not alter the direction or significance of the reported effect.

| **Characteristic** | **Amyloid Confirmation, N = 125†** | **PiB PET Imaging, N = 87†** | **Cognitive Testing, N = 103†** | **CSF Biomarkers, N = 91†** |
|---|---|---|---|---|
| Disease status | | | | |

| Characteristic | Amyloid Confirmation, N = 125† | PiB PET Imaging, N = 87† | Cognitive Testing, N = 103† | CSF Biomarkers, N = 91† |
|---|---|---|---|---|
| CU Aβ- | 79 (63%) | 55 (63%) | 72 (71%) | 54 (59%) |
| CU Aβ+ | 33 (26%) | 26 (30%) | 30 (29%) | 25 (27%) |
| AD dementia | 13 (10%) | 6 (6.8%) | 0 (0%) | 12 (13%) |
| Age at fecal sample, *years*[2] | 67.54 (6.46) | 67.46 (6.14) | 66.61 (6.38) | 67.20 (6.59) |
| Sex[1] | | | | |
| Female | 81 (65%) | 57 (66%) | 65 (63%) | 57 (63%) |
| Male | 44 (35%) | 30 (34%) | 38 (37%) | 34 (37%) |
| *APOE* genotype[1] | | | | |
| E2-E3 | 12 (9.9%) | 8 (9.2%) | 10 (10%) | 9 (10%) |
| E2-E4 | 2 (1.7%) | 0 | 2 (2.0%) | 2 (2.3%) |
| E3-E3 | 59 (49%) | 42 (48%) | 55 (56%) | 38 (44%) |
| E3-E4 | 41 (34%) | 32 (37%) | 31 (31%) | 32 (37%) |
| E4-E4 | 7 (5.8%) | 5 (5.7%) | 1 (1.0%) | 6 (6.9%) |
| Unknown | 4 | 0 | 4 | 4 |
| Education, *years*[2] | 16.17 (2.57) | 16.04 (2.56) | 16.40 (2.64) | 16.08 (2.48) |
| BMI, *kg/m*$^2$ | 27.72 (4.86) | 27.84 (5.05) | 27.64 (5.00) | 27.71 (4.59) |
| Fecal sample BSS | 3.87 (1.20) | 3.78 (1.18) | 3.90 (1.21) | 3.91 (1.18) |

†Mean (SD); n (%)

**Table S1. Characteristics of participants across all outcome measures.** *AD dementia*, Alzheimer's disease dementia, Aβ-positive; *BMI*, body mass index; *CU Aβ-*, cognitively unimpaired, Aβ-negative; *CU Aβ+*, cognitively unimpaired, Aβ-positive; *CSF*, cerebrospinal fluid; *PET*, positron emission tomography. Bristol Stool Scale (BSS) ranges from 1 (hard lumps) to 7 (liquid).

| **Characteristic** | **CU Aβ-, N = 55**[1] | **CU Aβ+, N = 26**[1] | **AD dementia, N = 6**[1] |
|---|---|---|---|
| Age at fecal sample, *years* | 66.49 (6.22) | 68.68 (5.96) | 71.14 (4.42) |
| Sex | | | |
| Female | 35 (64%) | 17 (65%) | 5 (83%) |
| Male | 20 (36%) | 9 (35%) | 1 (17%) |
| *APOE* genotype | | | |
| E2-E3 | 8 (15%) | 0 (0%) | 0 (0%) |
| E3-E3 | 31 (56%) | 10 (38%) | 1 (17%) |
| E3-E4 | 14 (25%) | 15 (58%) | 3 (50%) |
| E4-E4 | 2 (3.6%) | 1 (3.8%) | 2 (33%) |
| BMI, *kg/m²* | 27.88 (5.10) | 27.89 (5.07) | 27.27 (5.36) |
| Education, *years* | 16.13 (2.75) | 15.92 (2.31) | 15.83 (1.94) |
| Fecal sample BSS | 3.82 (1.12) | 3.73 (1.22) | 3.67 (1.63) |
| PiB PET cortical DVR | 1.05 (0.04) | 1.35 (0.24) | 1.67 (0.10) |

[1]Mean (SD); n (%)

**Table S2. Characteristics of participants with PiB PET neuroimaging.** Heading abbreviations are defined in table S1.

| Characteristic | CU Aβ-, N = 73[1] | CU Aβ+, N = 30[1] |
|---|---|---|
| Age at fecal sample, *years* | 65.58 (6.03) | 69.09 (6.62) |
| Sex | | |
| Female | 46 (63%) | 19 (63%) |
| Male | 27 (37%) | 11 (37%) |
| *APOE* genotype | | |
| E2-E3 | 9 (13%) | 1 (3.3%) |
| E2-E4 | 1 (1.4%) | 1 (3.3%) |
| E3-E3 | 42 (61%) | 13 (43%) |
| E3-E4 | 17 (25%) | 14 (47%) |
| E4-E4 | 0 (0%) | 1 (3.3%) |
| Unknown | 4 | 0 |
| BMI, $kg/m^2$ | 27.83 (5.06) | 27.16 (4.91) |
| Education, *years* | 16.44 (2.78) | 16.30 (2.29) |
| Fecal sample BSS | 3.95 (1.19) | 3.80 (1.27) |
| RAVLT: Trials 1-5, *words learned* | 52.65 (8.38) | 52.60 (11.63) |
| Unknown | 1 | 0 |
| RAVLT: Trial 7, *words recalled* | 10.71 (2.94) | 10.53 (3.77) |
| Unknown | 1 | 0 |
| TMT: Trial B Time, *seconds* | 58.60 (25.61) | 61.40 (28.14) |
| Unknown | 1 | 0 |

[1]Mean (SD); n (%)

**Table S3. Characteristics of participants with cognitive testing.** Heading abbreviations are defined in table S1.

| **Characteristic** | **CU Aβ-, N = 54**[1] | **CU Aβ+, N = 25**[1] | **AD dementia, N = 11**[1] |
|---|---|---|---|
| Age at fecal sample, *years* | 65.11 (6.22) | 68.21 (5.41) | 74.83 (4.89) |
| Sex | | | |
| Female | 33 (61%) | 16 (64%) | 7 (64%) |
| Male | 21 (39%) | 9 (36%) | 4 (36%) |
| *APOE* genotype | | | |
| E2-E3 | 8 (16%) | 1 (4.0%) | 0 (0%) |
| E2-E4 | 1 (2.0%) | 1 (4.0%) | 0 (0%) |
| E3-E3 | 27 (54%) | 9 (36%) | 2 (18%) |
| E3-E4 | 13 (26%) | 12 (48%) | 7 (64%) |
| E4-E4 | 1 (2.0%) | 2 (8.0%) | 2 (18%) |
| Unknown | 4 | 0 | 0 |
| BMI, $kg/m^2$ | 27.85 (4.46) | 28.02 (5.24) | 26.07 (3.72) |
| Fecal sample BSS | 3.89 (1.25) | 3.96 (0.89) | 3.82 (1.47) |
| Education, *years* | 16.44 (2.65) | 16.04 (2.03) | 14.36 (2.01) |
| $A\beta_{42}/A\beta_{40}$ | 0.07 (0.01) | 0.04 (0.01) | 0.03 (0.01) |
| $pTau_{181}/ A\beta_{42}$ | 0.02 (0.01) | 0.04 (0.02) | 0.08 (0.03) |
| tTau, *pg/mL* | 191.68 (56.17) | 251.57 (82.64) | 368.35 (147.34) |
| Neurogranin, *pg/mL* | 746.28 (254.28) | 952.21 (378.10) | 1,175.35 (546.73) |

| Characteristic | CU Aβ-, N = 54[1] | CU Aβ+, N = 25[1] | AD dementia, N = 11[1] |
|---|---|---|---|
| NFL, *pg/mL* | 90.87 (40.16) | 114.43 (53.88) | 192.13 (74.63) |
| GFAP, *ng/mL* | 9.83 (3.55) | 11.26 (3.31) | 14.97 (5.35) |
| YKL-40, *ng/mL* | 152.99 (51.57) | 166.41 (53.60) | 225.32 (93.60) |
| sTREM2, *ng/mL* | 7.91 (2.29) | 8.64 (2.58) | 9.94 (2.95) |
| α-Synuclein, *ng/mL* | 151.84 (58.04) | 167.62 (57.03) | 247.19 (105.01) |
| S100B, *ng/mL* | 1.10 (0.20) | 1.19 (0.27) | 1.31 (0.27) |

[1]Mean (SD); n (%)

**Table S4. Characteristics of participants with CSF biomarkers.** Heading abbreviations are defined in table S1.

| Reference Cohort | Predictor | Estimate ($\log_{10}$ μg/g) | *P*-value |
|---|---|---|---|
| | $calprotectin = disease\ status + sex$ | | |
| CU Aβ- | CU Aβ+ | 0.040 (-0.11, 0.20) | 0.59 |
| ***CU Aβ-*** | ***AD dementia*** | ***0.29 (0.080, 0.49)*** | ***0.010*** |
| ***CU Aβ+*** | ***AD dementia*** | ***0.24 (0.020, 0.47)*** | ***0.030*** |
| | $calprotectin = disease\ status + sex + age + BMI$ | | |
| CU Aβ- | CU Aβ+ | 0.00 (-0.16, 0.15) | 0.96 |
| CU Aβ- | AD dementia | 0.17 (-0.06, 0.39) | 0.15 |
| CU Aβ+ | AD dementia | 0.17 (-0.05, 0.39) | 0.13 |
| ***CU Aβ-*** | ***Age*** | ***0.020 (0.00, 0.030)*** | ***0.030*** |

**Table S5. Diagnosis and age effects on calprotectin levels.** Heading abbreviations are defined in table S1. Effects were tested using full participant cohort. β coefficients with significant *p*-values (multiple regression) are bolded and italicized. Disease status effects were significant before age was included in the model. While the effect of age was found significant over and above the effects of AD dementia status, its estimated magnitude was ten times smaller. No covariates had significant relationships with calprotectin level.

| Reference Cohort | Predictor | Estimate ($\log_{10}$ μg/g) | ***P*-value** |
|---|---|---|---|
| | $calprotectin = disease\ status$ | | |
| CU Aβ- | CU Aβ+ | 0.040 (-0.12, 0.20) | 0.61 |
| | $calprotectin = disease\ status + age + sex + BMI$ | | |
| ***CU Aβ-*** | ***Age*** | ***0.020 (0.00, 0.030)*** | ***0.040*** |
| CU Aβ- | CU Aβ+ | 0.00 (-0.16, 0.15) | 0.99 |

**Table S6. Age effects on calprotectin levels within cognitively unimpaired-only cohort.** Heading abbreviations are defined in tables S1 and S5. Effects were tested among cognitively unimpaired participants, comparing effects of age and CU Aβ+ status against the CU Aβ- reference cohort. β coefficients with significant *p*-values (multiple regression) are bolded and italicized. CU Aβ+ disease status effect was insignificant before adding model covariates. Age effect remained significant in full model. No covariates had significant relationships with calprotectin level.

| **Characteristic** | **Low, N = 69**[1] | **High, N = 10**[1] | ***P*-value**[2] |
|---|---|---|---|
| Calprotectin, $log_{10}(\mu g/g)$ | 1.00 (0.27) | 1.98 (0.24) | <0.0010 |
| Age at fecal sample, *years* | 66 (6) | 67 (7) | 0.40 |
| Cardiovascular disease | | | |
| Type 2 diabetes | 5 (7.2%) | 3 (30%) | 0.059 |
| Pre-diabetes | 13 (19%) | 4 (40%) | 0.20 |
| Microbiome-altering medications | | | |
| Proton pump inhibitor | 5 (7.2%) | 4 (40%) | 0.013 |
| SSRI antidepressant | 5 (7.2%) | 1 (10%) | 0.60 |
| Bulk-forming laxative | 6 (8.7%) | 1 (10%) | >0.90 |
| Diet | | | 0.70 |
| Vegetarian/vegan | 2 (1.4%) | 0 (0.00%) | |
| Meat/fish/poultry 1-2 days weekly | 8 (12%) | 0 (0.00%) | |
| Meat/fish/poultry 3+ days weekly | 59 (86%) | 10 (100%) | |
| Animal caretaking | | | |

| **Characteristic** | **Low, N = 69**[1] | **High, N = 10**[1] | ***P*-value**[2] |
|---|---|---|---|
| House pets | 33 (48%) | 6 (60%) | 0.50 |
| Other animals, e.g., farm animals | 8 (12%) | 1 (10%) | >0.90 |
| Early childhood microbial exposure | | | |
| Cesarian section birth | 3 (4.3%) | 0 (0.00%) | >0.90 |
| Breastfeeding in infanthood | 30 (54%) | 4 (57%) | >0.90 |

[1]Mean (SD); n (%)

[2]Wilcoxon rank sum test; Fisher's exact test

**Table S7. Lifestyle factors compared across low- and high-calprotectin CU Aβ- participants.** *SSRI*, selective serotonin reuptake inhibitor. Gut microbiome-altering medications were derived from the meta-analysis by Vich Vila et al. 2020; laxatives consumed by this cohort were identified using the Anatomical Therapeutic Chemical classification system.

| **Region** | **Estimate** | ***P*-value** | ***Q*-value** |
|---|---|---|---|
| $DVR = calprotectin + disease\ status + (calprotectin \times disease\ status) + age$ | | | |
| ANG | 0.244, (0.044, 0.445) | 0.020 | 0.030 |
| ACC | 0.197, (0.118, 0.275) | 8.66e-07 | 9.1e-06 |
| PCC | 0.568, (0.402, 0.733) | 2.0e-11 | 5.0e-10 |
| vmPFC | 0.194, (0.062, 0.326) | 0.004 | 0.008 |
| PCUN | 0.29, (0.168, 0.412) | 3.22e-06 | 2.54e-05 |
| SMG | 0.19, (0.004, 0.376) | 0.045 | 0.08 |
| MTG | 0.123, (0.011, 0.235) | 0.03 | 0.06 |
| STG | 0.124, (-0.083, 0.33) | 0.24 | 0.42 |

| | | | |
|---|---|---|---|
| global | 0.232, (0.141, 0.322) | 5.02e-07 | 6.32e-06 |
| $DVR = calprotectin + disease\ status + (calprotectin \times disease\ status) + age + sex + APOE + BMI$ | | | |
| ANG | 0.184, (-0.065, 0.433) | 0.15 | 0.43 |
| ACC | 0.131, (-0.018, 0.279) | 0.08 | 0.25 |
| PCC | 0.509, (0.278, 0.74) | 1.54e-05 | 0.0005 |
| vmPFC | 0.108, (-0.091, 0.307) | 0.29 | 0.64 |
| PCUN | 0.238, (0.067, 0.408) | 0.006 | 0.02 |
| SMG | 0.126, (-0.065, 0.318) | 0.20 | 0.50 |
| MTG | 0.067, (-0.079, 0.214) | 0.37 | 0.73 |
| STG | 0.062, (-0.142, 0.266) | 0.55 | 0.85 |
| global | 0.164, (0.025, 0.303) | 0.02 | 0.07 |

**Table S8. Calprotectin-by-AD effects on regional PiB cortical distribution volume ratio (DVR) sustained after controlling for age.** *ACC*, anterior cingulate cortex; *ANG*, angular gyrus; *MTG*, middle temporal gyrus; *PCC*, posterior cingulate cortex; *PCUN*, precuneus; *SMG*, supramarginal gyrus; *STG*, superior temporal gyrus; *vmPFC*, ventromedial prefrontal cortex; *global*, mean of all bilateral regions' cortical DVR. β coefficients were estimated using multiple regression ($q$, false discovery rate corrected per model group).

| Region | Estimate | *P*-value |
|---|---|---|
| ANG | -0.094, (-0.299, 0.112) | 0.371 |
| ACC | -0.081, (-0.304, 0.141) | 0.475 |
| PCC | -0.067, (-0.243, 0.109) | 0.457 |
| vmPFC | -0.077, (-0.306, 0.152) | 0.509 |

| | | |
|---|---|---|
| PCUN | -0.09, (-0.291, 0.112) | 0.383 |
| SMG | -0.082, (-0.253, 0.09) | 0.351 |
| MTG | -0.084, (-0.267, 0.099) | 0.368 |
| STG | -0.082, (-0.234, 0.07) | 0.29 |
| global | -0.075, (-0.256, 0.105) | 0.413 |

**Table S9. Calprotectin-by-CU Aβ+ interaction effects on regional PiB cortical DVR.** Region abbreviations are defined in table S8. β coefficients were estimated using the multiple regression equation $DVR = calprotectin + disease\ status + (calprotectin \times A\beta^{+}\ status) + age + sex + APOE + BMI$. Interactions effects were nonsignificant before adjusting for multiple comparisons.

| Region | Estimate | *P*-value | *Q*-value |
|---|---|---|---|
| ANG | -0.003, (-0.012, 0.005) | 0.435 | 0.726 |
| ACC | 0.001, (-0.009, 0.012) | 0.801 | 0.892 |
| PCC | 0.004, (-0.006, 0.014) | 0.413 | 0.726 |
| vmPFC | 0.002, (-0.009, 0.012) | 0.757 | 0.892 |
| PCUN | 0.001, (-0.009, 0.011) | 0.885 | 0.918 |
| SMG | -0.003, (-0.011, 0.005) | 0.475 | 0.726 |
| MTG | -0.003, (-0.012, 0.005) | 0.432 | 0.726 |
| STG | -0.004, (-0.012, 0.005) | 0.406 | 0.726 |
| global | -0.001, (-0.009, 0.008) | 0.892 | 0.918 |

**Table S10. Calprotectin-by-age interaction effects on regional PiB cortical DVR.** *ACC*, anterior cingulate cortex; *ANG*, angular gyrus; *MTG*, middle temporal gyrus; *PCC*, posterior cingulate cortex; *PCUN*, precuneus; *SMG*, supramarginal gyrus; *STG*, superior temporal gyrus;

*vmPFC*, ventromedial prefrontal cortex; *global*, mean of all bilateral regions' cortical DVR. β coefficients were estimated using multiple regression equation $DVR = calprotectin + (calprotectin \times age) + age$ ($q$, false discovery rate corrected).

| Biomarker | Estimate | *P*-value | *Q*-value |
|---|---|---|---|
| ***A$\beta_{42}$/A$\beta_{40}$*** | -0.001, (-0.009, 0.007) | 0.803 | 0.82 |
| ***pTau$_{181}$/A$\beta_{42}$*** | -0.004, (-0.016, 0.009) | 0.568 | 0.61 |
| tTau, *pg/mL* | -51.057, (-98.961, -3.153) | 0.037 | 0.08 |
| ***NFL, pg/mL*** | -22.322, (-44.855, 0.211) | 0.052 | 0.11 |
| Neurogranin, *pg/mL* | -192.257, (-394.375, 9.862) | 0.062 | 0.12 |
| α-Synuclein, *ng/mL* | -43.583, (-82.352, -4.814) | 0.028 | 0.07 |
| sTREM2, *ng/mL* | -1.051, (-2.487, 0.385) | 0.152 | 0.23 |
| YKL-40, *ng/mL* | -52.253, (-80.795, -23.711) | 0 | 0 |
| GFAP, *ng/mL* | -1.416, (-3.733, 0.902) | 0.231 | 0.3 |
| S100B, *ng/mL* | -0.112, (-0.266, 0.042) | 0.155 | 0.23 |

**Table S11. Calprotectin relationships with CSF biomarkers, controlled for age and disease status.** β coefficients were estimated using multiple regression $DVR = calprotectin + age + disease\ status$ ($q$, false discovery rate corrected).
